# Attention and interoception alter perceptual and neural pain signatures

**DOI:** 10.1101/2023.11.04.23298049

**Authors:** Maria Niedernhuber, Joaquim Streicher, Bigna Lenggenhager, Tristan A. Bekinschtein

**Affiliations:** Consciousness and Cognition Lab, Department of Psychology, University of Cambridge; Department of Psychology, University of Zurich; Department of Psychology, University of Konstanz, Germany

**Keywords:** pain, consciousness, EEG, power, CRPS

## Abstract

Fluctuations of chronic pain levels are determined by a complex interplay of cognitive, emotional and perceptual variables. We introduce a pain tracking platform composed of wearable neurotechnology and a smartphone application to measure and predict chronic pain levels. Our method measures, dynamically, pain strength, phenomenal and neural time series collected with an online tool and low-density EEG. Here we used data from a single participant who performed an attention task at home for a period of 20 days to investigate the role of attention to different bodily systems in chronic pain. Our results show a relationship between emotions and pain strength while allocating attention to the heartbeat, the breathing, the affected or the unaffected limb. We found that pain was maximal when attending to the affected limb, and decreased when the participant focused on his breathing or his heartbeat. These results provide interesting insights regarding the role of attention to interoceptive signals in chronic pain. We found power changes in the delta, theta, alpha and beta (but not in the gamma) band between the four attention conditions. However, there was no reliable association of these changes to pain intensity ratings. Theta power was higher when attention was directed to the unaffected limb compared to the others. Further, the pain ratings, when attending to unaffected limb, were associated with alpha and theta power band changes. Overall, we demonstrate that our neurophysiology and experience tracking platform can capture how body attention allocation alters the dynamics of subjective measures and its neural correlates. This research approach is proof of concept for the development of personalized clinical assessment tools and a testbed for behavioural, subjective and biomarkers characterization.

## Introduction

Chronic pain is a pressing public health issue which drastically impairs quality of life, affecting approximately one third of the world’s population at some point during their lives (Fayaz et al., 2016). In recent decades, scientific consensus emerged that chronic pain is not a direct readout of a nociceptive event but rather a conscious experience shaped by complex interactions of emotional and cognitive processes (Bushnell et al., 2013; Colloca et al., 2013; Lee et al., 2009; Wiech, 2016; Wiech et al., 2008). A key challenge is now to disentangle the complex interplay of internal and external psychological factors influencing pain levels to develop informed interventions for pain relief and ameliorate wellbeing.

In this study, we examined whether manipulations of attention to different bodily systems influence chronic pain (pain strength) and pain-related (emotion valence, body perception) perception. Attention, emotion, and interoception, which is the sense of one’s internal bodily states (Craig, 2002), have been shown to be important drivers and modulators of chronic pain (Bantick et al., 2002; Di Lernia, Serino, & Riva, 2016). Moreover, chronic pain disrupts the interoceptive system which processes signals arising from within the body (Di Lernia, Serino, & Riva, 2016), and individuals living with chronic pain show impairments across various interoceptive tasks such as interoceptive accuracy in heartbeat detection paradigms (Di Lernia et al., 2020; Di Lernia, Serino, Cipresso, et al., 2016; Di Lernia, Serino, & Riva, 2016; Solcà et al., 2020). Interestingly, exteroceptive display of interoceptive signals has been shown to lead to pain relief (Solcà et al., 2018). Interoception is also often considered crucial for emotional processing (Seth, 2013; Seth & Critchley, 2013), which is itself impaired in chronic pain (Bushnell et al., 2013; Geha et al., 2008). In line with this, some studies support an interplay of interoceptive and emotional dysfunction in chronic pain (Borg et al., 2018). Abundant evidence also describes the powerful influence of attention on chronic pain. Across different studies, pain levels decrease when attention is diverted away from the perceived origin of pain and increases when perceived tissue damage is attended to (Bantick et al., 2002; Defina et al., 2021; Eccleston, 1995). However, previous work concentrated on the influence of attention on pain in the exteroceptive domain, e.g., using auditory or visual stimuli (Wiech et al., 2008). An interesting avenue of research is interoceptive attention, which is a process in which attention is directed to signals arising from within the body (X. Wang et al., 2019). Indeed, there is some evidence suggesting that breathing exercises lead to pain relief across various chronic pain conditions (Mehling et al., 2005; Park et al., 2013; H. Wang et al., 2023). Similarly, exteroceptive presentation of cardiac signals has shown to relieve chronic pain (Solcà et al., 2018). Considering that interoceptive dysfunction typically accompanies CRPS, the question arises whether attention to interoceptive function leads to pain relief. Furthermore, many clinical pain disorders are associated with disorders of body representation and attention to the body as compared to distraction has been suggested to be beneficial for normalizing such conditions.

This work is part of a larger study that uses Complex Regional Pain Syndrome (CRPS) as a model to assess the complex interplay between subjective and objective measures, between experience and neural biomarkers (Jachs et al., 2022). CRPS is a chronic pain condition with a devastating impact on quality of life. It manifests as a result of a minor injury which leads to disproportionate chronic pain commonly linked to inflammatory responses. A hallmark of CRPS is that individuals experience a pronounced variety of concomitant cognitive, emotional and perceptual changes such as distortions of body perception, depression, and hemispheric inattention (Kuttikat et al., 2016). Here, in 20 sessions, an individual with unilateral CRPS in his left leg was asked to direct his attention either to his breathing, heartbeat, painful or unaffected leg for several minutes while we recorded EEG. After every condition, we collected the participant’s ratings using temporal experience tracing (TET): the participant was asked to draw how he felt during the attention task session on a grid, the x-axis corresponding to time and the y-axis corresponding to the intensity of various phenomenal dimensions (cognition, emotion, body perception and pain intensity) (see Figure 1). In line with past work (Wiech, 2016), we determined whether attention to pain enhances its intensity. As per the pre-registration in OSF (https://osf.io/6wu2k/), our prediction was that negative emotions and distortions of body perception would increase with reported pain intensity. We also tested the hypothesis whether the powerful crosstalk between attention and interoception can be exploited for pain relief. Here we hypothesised that attention to interoceptive processes (breathing, heartbeat) decreases pain strength. We also aimed to test for differences in power spectral density when attention is directed either to the painful leg, unaffected leg, one’s breathing or heartbeat. To further explore the relationship between pain intensity and other phenomenal experience dimensions (negative emotion, positive emotion, perception of bodily changes, lack of ownership), we performed association analyses across all four conditions, and correlation analysis between EEG power bands and pain strength as a first characterization of this single case in neurophenomenology of chronic pain.

**Figure 1.**
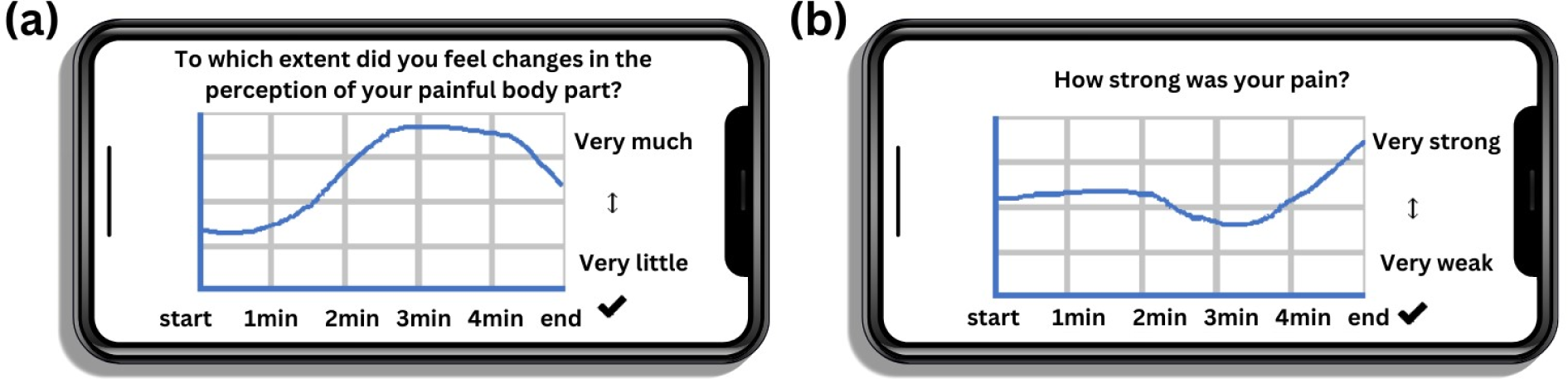
A new interface to report pain intensity and measure pain-related experience dimensions. *Note* Participants can use their laptop or their smartphone to report their experience during the attention task on a grid mapping experience intensity along a temporal dimension. Here two examples are given: (a) changes in body perception: *To which extent did you feel changes in the perception of your painful body part?* and (b) pain intensity: *How strong was your pain?*.

## Methods

### Participant

We tested our hypotheses in a man with unilateral CRPS aged 41-45 years old. Our participant first developed symptoms consistent with CRPS in 2008 and was later diagnosed with CRPS in 2012. His symptoms were localised in the left leg, including toes, foot, ankle, calf, shin, knee, front and side of the thigh. At the time of the experiment, he was taking Pregabalin (300-600mg in the morning and at night), Tramadol (100-200mg in the morning and at night), Codeine occasionally (60-240mg per day, when taken) and THC oil (40-50mg per day at noon and in the evening). Data was collected in November and December 2022.

### Software development

We developed a mobile application which collects phenomenal time series of chronic pain and related sensations across various domains. Temporal experience traces (TET) track the time course of the intensity of a specific component of (pain) experience and can be seen as a temporally extended version of a Likert-type point scale rating, i.e., a dimension intensity is measured at each instant t, resulting in a temporal curve. The traces of experience developed for this project are an extension of the methods developed previously at the CCC-Lab (Jachs et al., 2022). The application records participative dimensions linked to chronic pain such as participant’s perceived intensity and characteristics of their pain, the evolution of their emotional states, as well as their body perception. This version of the TET method allows us to track participant’s experience across all these dimensions, and to align these phenomenal time series to cortical data measured with low-density electroencephalography (EEG).

### Study procedure and data collection

The experiment took place on 20 testing days at the participant’s home. Every day, the participant completed a short task testing the influence of attention to the body while low-density EEG was recorded. The participant first attended a virtual training session guided by the experimenter in which he learned how to use EEG equipment and perform retrospective tracking of phenomenological experience components. During the task, the participant was asked to direct his attention to either his painful leg, his unaffected leg, to his heartbeat, or his breathing for 5 mins each while recording EEG. Conditions were presented in a randomised order. Each session was followed by the TET report, capturing the intensity of attention to the target body process, distortions of body perception, emotional states, and pain strength. For each component of experience, the participant was asked to indicate the intensity of his experience using a grid mapping experience ratings on an ordinal intensity scale from “very low” to “very strong” on time (1-5 min) (Figure 1). Finally, the participant was prompted to indicate which descriptors applied to his experience for every rating. Note that while a mobile app was developed, this participant reported his phenomenal experience using a web version of the app, designed using JATOS (Lange et al., 2015). The attention task is identical, but the participant was able to use his laptop instead of his phone as per this participant’s preference. The study received ethical approval from the Cambridge Psychology Research Ethics Committee.

We included the following items, with additional descriptors: 1. Pain strength (“very weak” to “very strong”): burning, stabbing, numbing, pins-and-needles, or throbbing. 2. Positive emotion (“Very good” to “normal”): happy, peaceful, relaxed, calm. 3. Negative emotion (“Very bad” to “normal”): sad, anxious, irritable, angry. 4. Body perception (“Strong changes” to “no changes): different weight, different size, different temperature, feeling alien, moves without control. 5. Lack of ownership over the painful limb (”very much” to “not at all”). 6. Attention to pain (“very much” to “not at all”). 7. Attention to task (“very much” to “not at all”). Collected data was stored on a cloud server until it was analysed. The data workflow is summarized in Figure 2.

**Figure 2.**
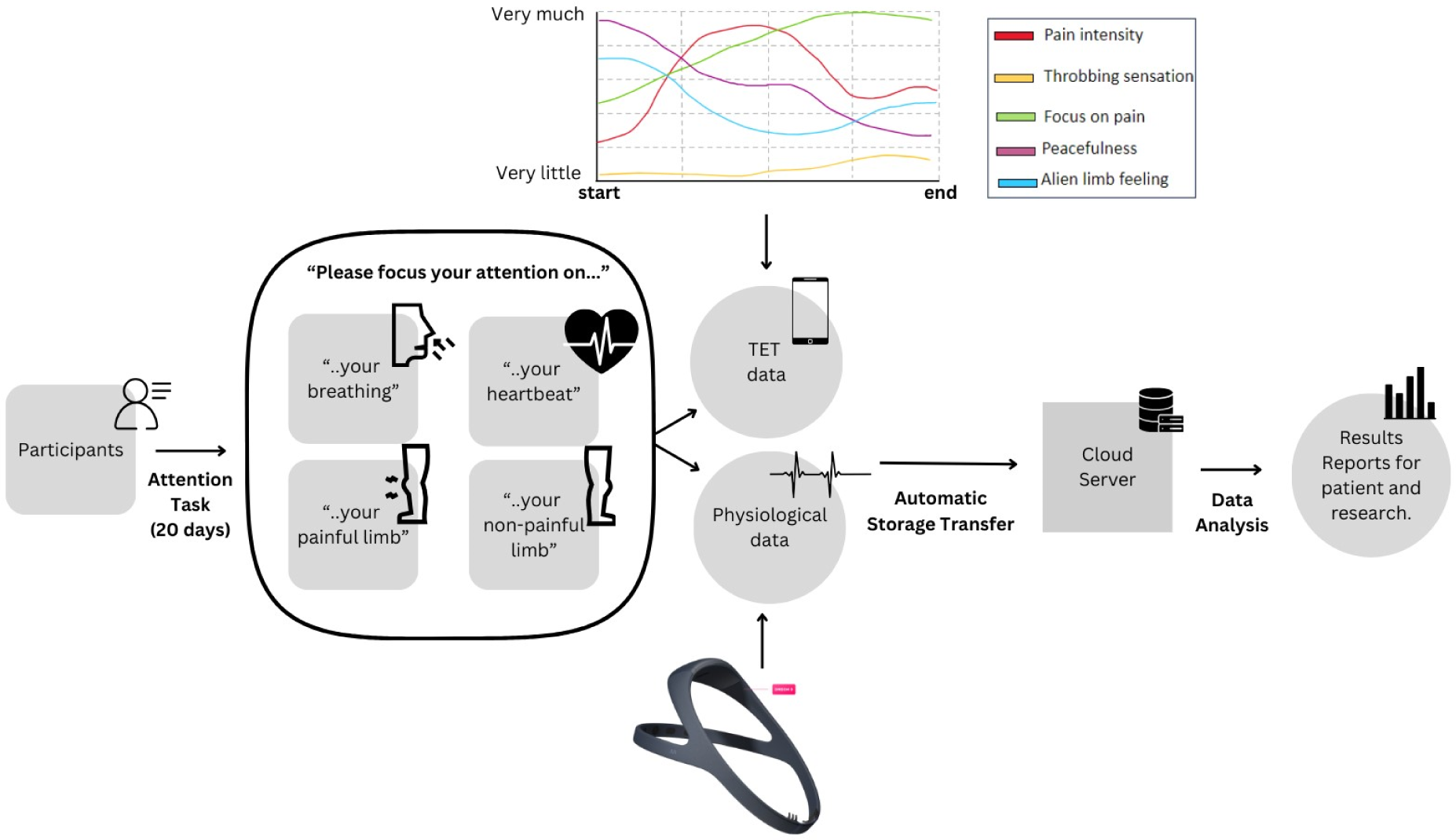
Data workflow. *Note.* The participant performed the attention task every day for 20 days, alternatively focusing on his painful limb, unaffected limb, breathing, or heartbeat while neurophysiological data was recorded using low-density EEG. The participant retrospectively rated his phenomenal experience (pain intensity, emotional states, body perception and ownership, attention capacity) after each 5min sub-session using the interface that was developed for the study, resulting in phenomenal time series than can be further analysed. Data is stored on a cloud server until it is analysed.

### Data preprocessing

We used low-density wearable EEG headbands with seven channels (Dreem, FDA Class II Medical Device) for cortical data collection. EEG data were preprocessed in Matlab R2021b using EEGLAB and custom-made functions. Initially, we applied a high-pass filter at 1 Hz to the data. We segmented the continuous data into epochs of 4 seconds. Epochs were rejected using a semi-automated preprocessing pipeline in which segments were rejected if data exceeded +-350 µV, a slope maximum at 0.3, or a slope r at 0.2. Phenomenal time series were imported in Javascript using Visual Studio Code Version 1.82.1 and processed in R Version 4.2.3 using RStudio Version 2023.06.0+421. Due to EEG recording errors, we retained 15 EEG datasets for further analysis.

### Data analysis

#### Power analysis

We tested for differences in EEG signal power between the four experimental conditions included in the attention task using Matlab 2021b and the EEGLAB toolbox (Delorme & Makeig, 2004). Based on EEG data quality after preprocessing, we included 15 runs of the attention task performed by the participant across 2 months. For each dataset, EEG were bandpass-filtered for each frequency range of interest using a Butterworth filter (alpha: 8-12 Hz, beta: 13-30 Hz, theta: 4-8 Hz, delta: 0.5-4 Hz, low gamma: 30:40 Hz). For each resulting dataset, the data were first Hilbert-transformed by calculating *V*(*t*) = *v*(*t*) + *i* ∗ *v*′(*t*) where a vector *V*(*t*) has a real part *v*(*t*) and an imaginary part *i* ∗ *v*′(*t*). To calculate the envelope, the analytic signal of Hilbert transform was power-transformed. We averaged the output per channel across the frequency range of interest, while at the same time retaining trials. We calculated the mean signal across 5 bins (each representing 1 min of testing time) for every dataset, resulting in 75 data points per condition. Finally, we performed a Friedman ANOVA to test for differences in EEG signal power between experimental conditions per frequency band of interest.

#### Phenomenal ratings analysis

We recorded phenomenal time series measuring body perception, body ownership, positive emotion, negative emotion, attention to task and pain strength after every experimental block lasting for 5 min. Time series were divided up in five segments lasting 1 min each, and averaged across each bin. We pooled score averages from 15 sessions (those for which good quality EEG data was available) included in our original 20 sessions dataset, leading to 75 data points per condition. For each rating type, we performed a Friedman test which revealed whether phenomenal ratings differ between the four experimental conditions (attention focused on painful limb, unaffected limb, breathing, heartbeat).

Note that the ownership dimension was missing a session, resulting in a 14-day dataset (i.e., 70 data points). The positive emotion dimension was missing two sub-sessions, namely the pain condition from one session and the heartbeat condition from another session. This absence of data might either be due to a problem caused by the software we used to collect data or to an error of the participant.

#### Correlation analysis

To investigate the relationship between pain strength and other phenomenal experience dimensions (negative emotion, positive emotion, perception of bodily changes, lack of body ownership), we ran a Kendall rank correlation on the phenomenal time series across sessions, per condition. Note that the corresponding data from the pain strength time series was excluded when the correlation was run with dimensions missing a session of data. To investigate the relationship between neural data and reported pain intensity, we also performed a Kendall rank correlation analysis between pain strength and the extracted power bands (alpha, beta, theta, delta, low gamma), resulting in 36 correlation analyses in total. A false discovery rate (FDR) multiple comparison correction was applied to all p values obtained from these analyses.

#### Preregistration

All our hypotheses and predictions, as well as the data analysis procedure, were preregistered on OSF (https://osf.io/6wu2k/).

## Results

### Attention to the body influences pain, emotion and cognition

Here we provide proof-of-concept that attention to the body influences chronic pain strength, characterising the influence of attention to exteroceptive and interoceptive processes on pain perception. We used a one-way repeated-measures ANOVA to show that attention to one’s body influences pain intensity (*X̃*^2^(3)=61, p<0.001). In line with previous work (Bantick et al., 2002; Wiech et al., 2008), we show that pain intensity was higher when the participant attended to the painful limb than the unaffected limb (Z=826, p=0.01). Importantly, our findings demonstrate that attention to interoceptive processes reduced pain in this patient. Pain intensity was lower when the participant concentrated on his heartbeat than on the painful limb (Z=504, p<0.001) or the unaffected limb (Z=984, p=0.12). Supporting the idea that attention to interoceptive processes reduces pain intensity, we show that pain intensity decreased when the participant attended to his own breathing relative to the painful limb (Z=216, p<0.001) and the unaffected limb (Z=294, p<0.001). We also found that attention to one’s breathing led to a stronger pain reduction than attention to one’s heartbeat (Z=630, p<0.001). Results are summarized in Figure 3.

**Figure 3.**
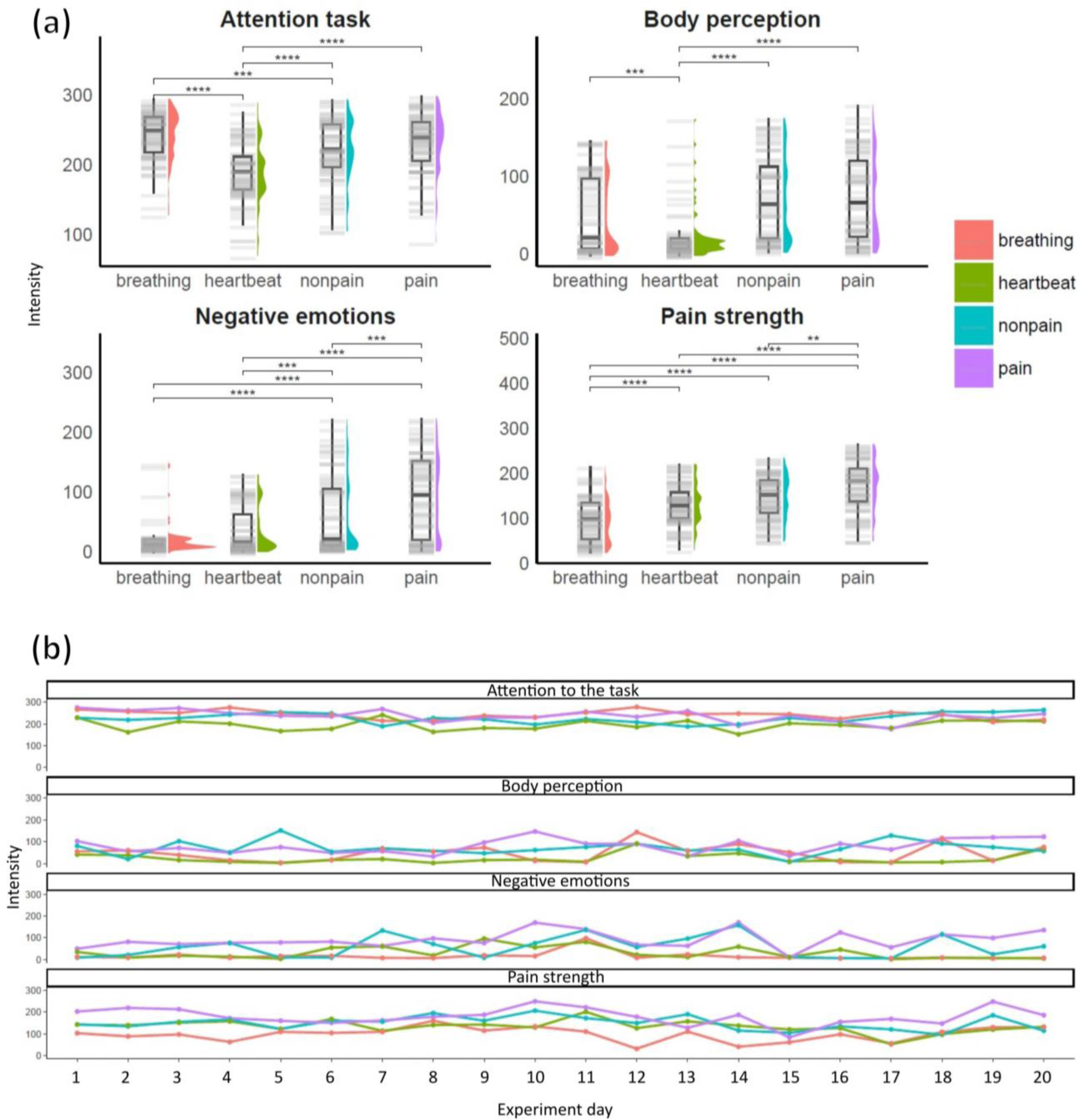
Attention modulates pain strength, body perception, and negative emotions. *Note.* **(a)** Panels show distributions of temporal experience traces (TET) averaged across 1 min bins for each of the 15 sessions. For each condition, power distributions are shown as a violin plot adjacent to a box plot and a barcode plot, and reliable differences between conditions resulting from post-hoc testing were included. We found an impact of the attentional focus (breathing, heartbeat, non-painful limb, painful limb) on all experience dimensions (negative emotions, body perception, and pain strength). Crucially, pain strength differed across all conditions, being the strongest when the participant focus his attention on his painful limb, and the weakest when he focused his attention on his breathing. **(b)** Ratings intensity distribution across the 20-day period. The four conditions (attention either focused on breathing, heartbeat, painful limb, or non-painful limb) are indicated following the same colour code as in part (a). The corresponding intensity is displayed per dimension (attention to task, body perception, negative emotion, pain strength) for all 20 sessions.

Moreover, we found that attention to the body influenced the extent to which the participant was able to focus on the task (*X̃*^2^(3)=60.65, p<0.001). The participant was less able to focus on his heartbeat than the painful leg (Z=341, p<0.001), unaffected leg (Z=578, p<0.001) or his breathing (Z=2627, p<0.001). We identified no differences in the participant’s ability to concentrate on his painful leg and either the unaffected leg (Z=1112, p=0.59) or breathing (Z=1714, p=0.77). The participant was able to focus better on his breathing than his unaffected leg (Z=2114, p=0.002) but we did not reveal any differences between attention to breathing and the painful leg (Z=1714, p=0.77).

Our data show that negative emotions differed between the four experimental conditions (*X̃*^2^(3)=51.78, p<0.001). Possibly due to an increase in pain strength, the participant felt worse when he attended to his painful leg than his other leg (Z=747, p=0.002), heartbeat (Z=260, p<0.001), or breathing (Z=204, p<0.001). The participant felt better when concentrating on his unaffected limb than his heartbeat (Z=760, p=0.003) or breathing (Z=606, p<0.001). Interestingly, attention to breathing increased positive emotions more than to the heartbeat (Z=889, p=0.03), suggesting differences between both interoceptive functions in the extent to which they influence perception.

Finally, we found that extent to which the participant reported shifts in body perception was different between experimental conditions (*X̃*^2^(3)=42, p<0.001). Interestingly, attention to the painful body part did not produce stronger distortions of body perception that the unaffected body part (Z=1216, p=1). However, the participant experienced fewer changes in body perception when he attended to his heartbeat than his painful leg (Z=185, p<0.001) or unaffected leg (Z=172, p<0.001). Body perception was more altered when the participant concentrated on his breathing than his heartbeat (Z=1885, p=0.001). We did not identify any differences in body perception between attention to breathing and the painful leg (Z=815, p=0.07) or the unaffected leg (Z=833, p=0.1).

### Neural power bands differs between attention allocation conditions

We set out to determine whether there are differences in power spectral density when the participant directs his attention to his breathing, heartbeat, his painful limb or his unaffected hand. In a series of one-way repeated measures ANOVAs, we revealed differences between conditions in the alpha (F(2.25137.05)=6.12, p=0.01, eta2[g]=0.06), beta (F(2.42133.07)=4.6, p=0.04, eta2[g]=0.06), theta (F(1.7887.42)=17.5, p<0.001, eta2[g]=0.12), and delta band (F(2.19118.51)=7.95, p=0.002, eta2[g]=0.06) but not in the gamma band (F(3156)=3.54, p=0.08, eta2[g]=0.04). Although power spectral measures differentiated experimental conditions, we did not observe that power changes mapped onto pain relief patterns observed in ratings. Theta power was reduced when attention was focused on the painful limb relative to the unaffected limb (t(49=7.78), p<0.001). Conversely, theta power was higher when the participant paid attention to his unaffected limb relative to his breathing (t(49=-3.57), p=0.02) or his heartbeat (t(49=-5.6), p<0.001). There were no theta power differences between attention to breathing and heartbeat (t(49=0.99), p=1) as well as pain and breathing (t(49=1.23), p=1) or heartbeat (t(49=0.81), p=1).

We found that beta power was higher when attention was directed to breathing than to heartbeat (t(55=3.84), p=0.01). There were no beta power differences between conditions in which the participant attended to his breathing versus the unaffected limb (t(55=0.67), p=1) or the painful limb (t(55=1.38), p=1). Likewise, beta power did not differ between conditions in which the participant focused his attention on his heartbeat versus his unaffected limb (t(55=-2.4), p=0.48) or painful limb (t(55=-2.73), p=0.2). Beta power was the same when attention to the unaffected versus painful limb was compared (t(55=0.13), p=1).

In the alpha band, we found that power increased when attention is paid to the unaffected limb than to one’s heartbeat (t(61=-4.45), p<0.001) but not one’s breathing (t(61=-3.1), p=0.07). We did not identify any alpha power differences between both interoceptive conditions (t(61=-0.68), p=1). Alpha power did not differ when attention was directed to one’s painful limb and either breathing (t(61=-2.35), p=0.53), heartbeat (t(61=-1.56), p=1) or the unaffected limb (t(61=1.78), p=1).

In the delta frequency range, power decreases when the participant attends to his heartbeat relative to the painful limb (t(54=-3.98), p=0.004) or unaffected limb (t(54=-5.72), p<0.001). We also found higher delta power during attention to breathing than heartbeat (t(54=3.43), p=0.03). We did not find delta power differences between attention to breathing and the unaffected limb (t(54=-2.2), p=0.76) or painful limb (t(54=-0.95), p=1). Likewise, a comparison of delta power measured during attention to the unaffected versus painful limb yielded no reliable results (t(54=1.19), p=1). Results are summarized in Figure 4.

**Figure 4.**
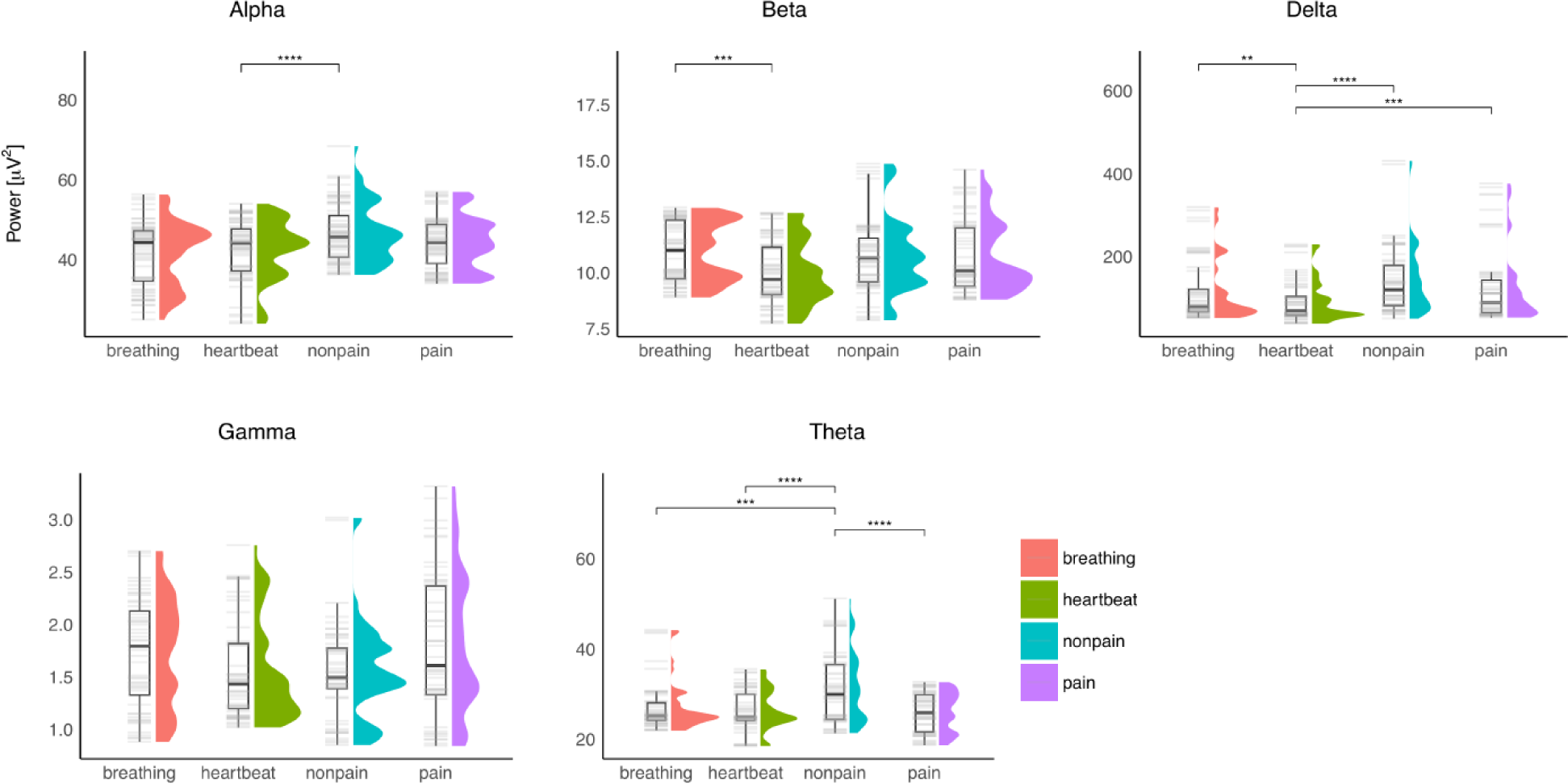
Attention modulates alpha, beta, delta and theta power bands, but not gamma. *Note.* Panels show distributions of time series averages obtained across 1 min bins for each of the 15 sessions. For each condition, power distributions are shown as a violin plot adjacent to a box plot and a barcode plot, and reliable differences between conditions resulting from post-hoc testing were included. We found a significant difference between at least two conditions (attention allocated either to participant’s breathing, heartbeat, non-painful limb, or painful limb) for all power bands except gamma.

### Relationship between pain strength and emotional states

As predicted, our results supported that experiencing negative emotion is positively correlated with pain intensity (Figure 5). More specifically, after FDR correction, the correlation between pain intensity and negative emotion was reliable when the participant were focusing his attention on his painful limb (tau=0.43, p<0.001) and on his heartbeat (tau=0.32, p<0.001). The correlations for the unaffected limb (tau=0.15, p=0.12) and breathing (tau=0.03, p=0.81) conditions were not reliable. On the other hand, the results from the correlation between pain intensity and positive emotion produced opposite results depending on the condition: there was a reliable positive correlation in the heartbeat condition (tau=0.20, p=0.04) but a reliable negative correlation in the breathing condition (tau=-0.27, p<0.01). Results for the painful limb (tau=-0.19, p=0.06) and unaffected limb (tau=-0.07, p=0.57) conditions were not reliable after FDR multiple comparison correction.

**Figure 5.**
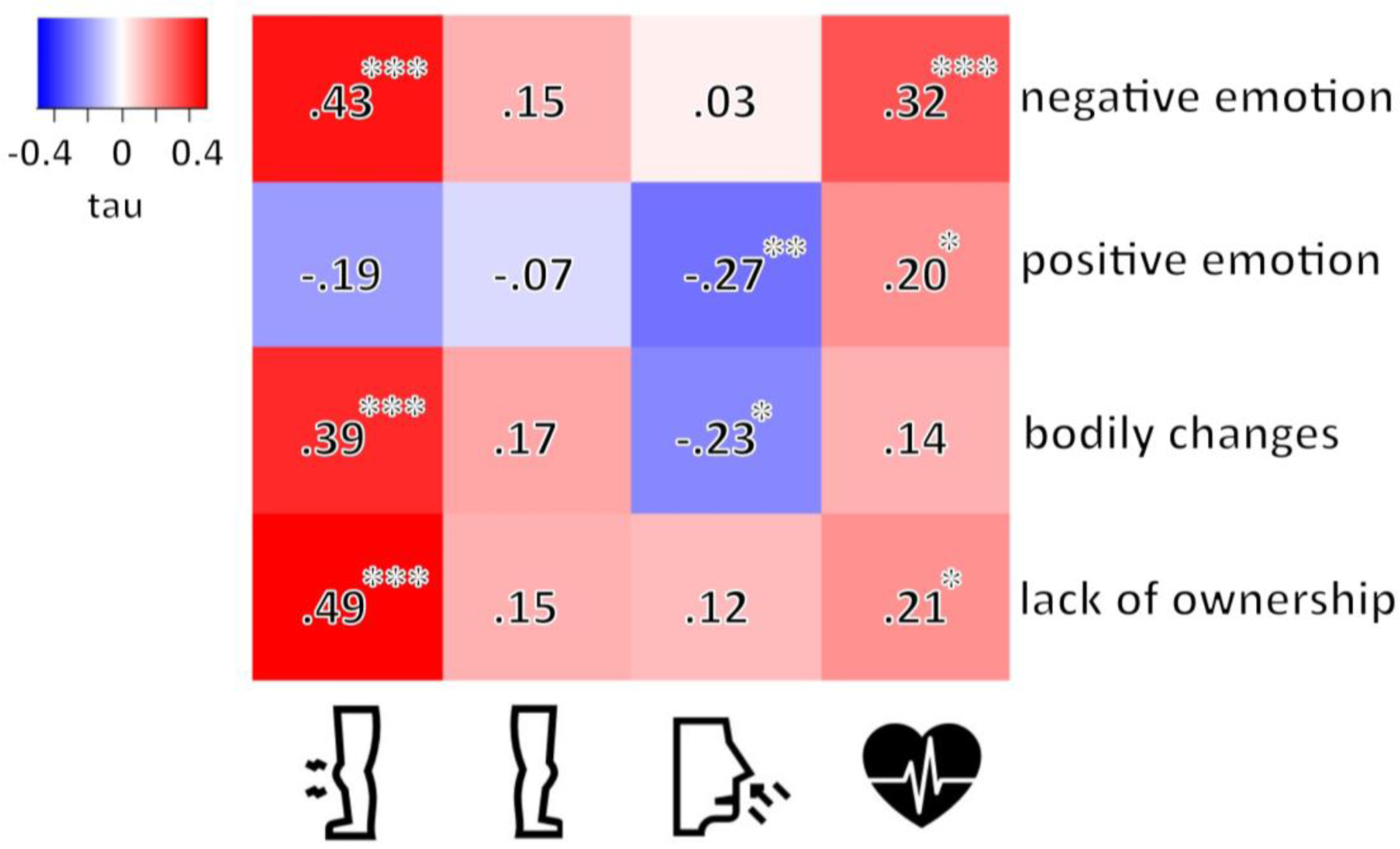
Attention modulates the relationship between pain strength and emotion, bodily changes, and limb ownership experience dimension. *Note.* Pain strength is correlated with negative emotions, the perception of bodily changes, and the sensation of lack of ownership when attention is allocated to the painful limb but not when attention is allocated to a non-painful body part or one’s own breathing. Heatmap summarizing the results from the Kendall rank correlation analyses between pain strength and other phenomenal time series, across all four conditions. Tau values are indicated. \**p<.05, **p<.01, **\*\*\***p<.001* after FDR correction. Crucially, we found that pain strength was reliably correlated with pain-related dimensions (negative emotions, bodily changes, lack of ownership) when the participant focused on his painful limb, but not when he focused on his non-painful limb or breathing. Interestingly, pain strength was negatively correlated with the perception of positive emotions and bodily changes when the participant focused on his breathing. Somewhat surprisingly, pain strength was positively correlated with the perception of negative emotions, lack of ownership, and positive emotions, when attention was allocated to the heartbeat.

### Relationship between pain strength and body perception

Changes in body perception were positively correlated with pain intensity when the participant was focusing on his painful limb (tau=0.39, p<0.001) but, interestingly, negatively correlated when the participant was focusing on his breathing (tau=-0.23, p=0.02). Results for the other conditions (unaffected limb: tau=0.17, p=0.08; heartbeat: tau=0.14, p=0.13) were not reliable. The sensation of a lack of ownership was positively correlated with pain intensity both for the painful limb (tau=0.49, p<0.001) and heartbeat (heartbeat: tau=0.21, p=0.04). Results for the two other conditions were not reliable (unaffected limb: tau=0.15, p=0.13; breathing: tau=0.12, p=0.20). All results were summarized as a heatmap, see Figure 5.

### Relationship between pain strength and neurophysiological activity

Most results from the correlation analysis between pain strength and power bands’ intensity were not reliable after the FDR correction was applied. There were two reliable positive correlations for the unaffected condition, for the alpha (tau=0.25, p=0.01) and theta (tau=0.22, p=0.03) power bands. All results were summarized as a heatmap, see Figure 6.

**Figure 6.**
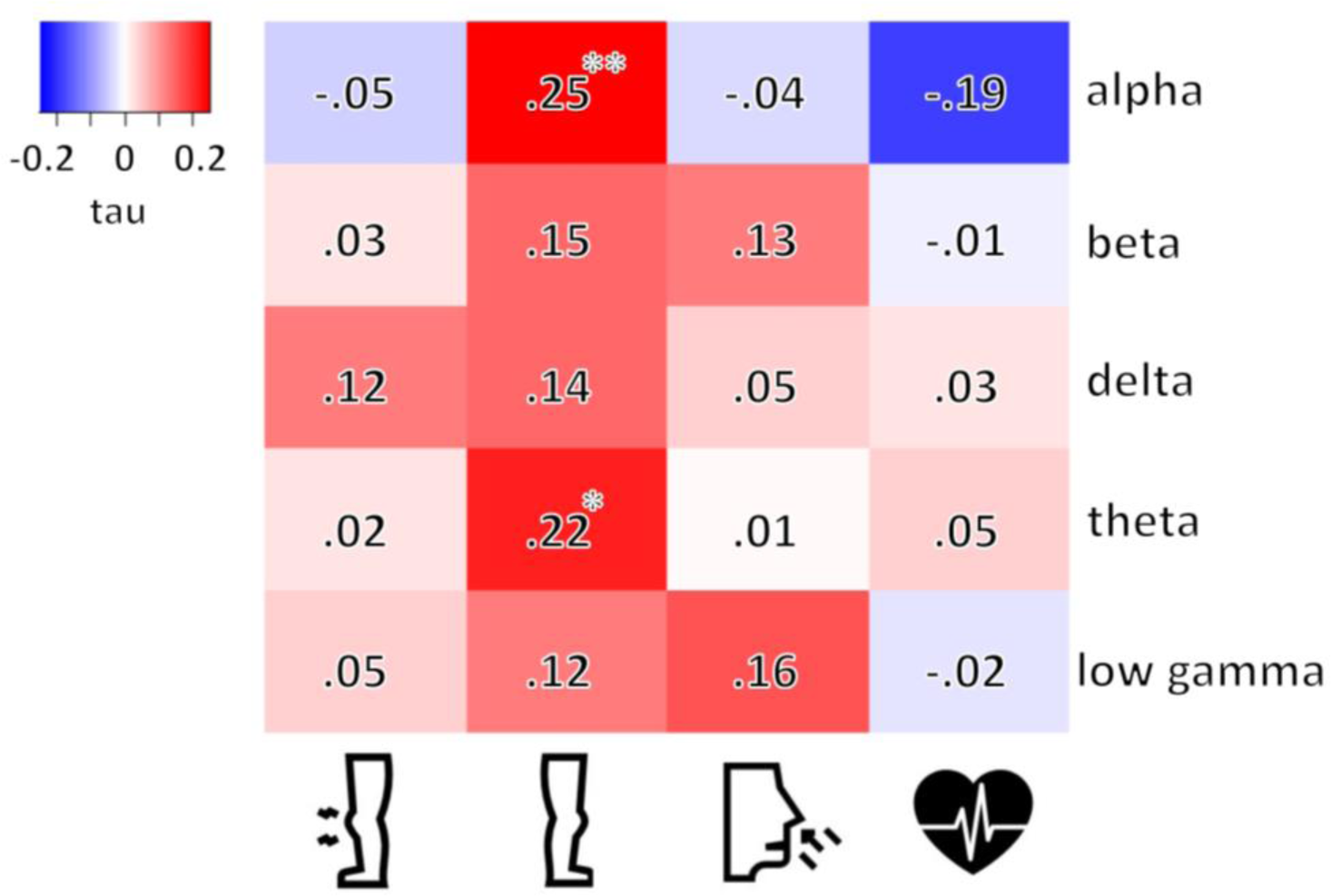
Pain strength associations to neural power bands depends on attention allocation. *Note.* Heatmap summarizing the results from the Kendall rank correlation analyses between pain strength and neural time series, across all four conditions. Tau values are indicated. *\*p<.05, **p<.01* after FDR correction. Somewhat counter-intuitively, the only reliable correlations were obtained in the non-painful limb condition, with alpha and theta intensity positively correlating with pain strength. We found no reliable correlation for the other conditions.

## Discussion

### Interoceptive attention produces hypoalgesic effects

In a case study combining systematic longitudinal cognitive testing and neurophysiological measures sampled with EEG, we determined how attention to the body influences the intensity of chronic pain as well as emotion and body perception. Supporting previous studies (Wiech et al., 2008), we found pain intensity increases when attention is paid to the site of chronic pain, and decreases when the unaffected body part is attended. Importantly, our results provide proof-of-principle that attention might induce pain relief when the focus is on one’s own breathing or heartbeat. Previous work shows a benefit of breathing exercises typically involving instructed slow breathing for pain relief (Jafari et al., 2016, 2020; H. Wang et al., 2023; Zunhammer et al., 2013) across a variety of chronic pain conditions (Mehling et al., 2005; Park et al., 2013; H. Wang et al., 2023). Importantly, our study directed the participant’s focus solely towards his breathing, akin to his attention on the affected or unaffected limb, without specific guidelines on breathing techniques. Considering that breathing exercises for pain relief involve interoceptive awareness, our findings suggest the potential analgesic effect of focused interoceptive attention alone. While previous research has indicated that both diversion from the source of pain and the anticipation of pain relief may contribute to hypoalgesia during the breathing exercise (Wiech et al., 2008), it is unlikely that these factors fully account for the pain relief experienced during breathing. Notably, our results revealed that the pain-alleviating impact of focused breathing surpassed that of shifting attention to the unaffected limb or even one’s heartbeat. Respiratory-induced hypoalgesia has been linked to physiological variables such as baroceptor (Reyes del Paso et al., 2015) or vagal nerve stimulation (Botha et al., 2015; Busch et al., 2013). Beyond interoceptive attention, those variables might explain the additional pain relief during attention to the breath.

In addition to breathing, attention to cardiac activity led to pain relief relative to attention to either the painful or unaffected body part, strengthening the evidence of the attention to interoceptive signals as a pain modulator. It has been shown in CRPS a reduction in the ability to detect one’s own heartbeat (in a heartbeat counting task) as well as depressed heartbeat-evoked potential amplitudes (Solcà et al., 2020). We speculate that attention to cardiac activity might modulate pain perception by enhancing interoceptive signalling.

### Attention to breathing differs from attention to heartbeat in several aspects of experience

Interestingly, the two interoceptive conditions differed in the extent to which they led to pain relief. We found that pain relief was stronger when the participant concentrated on his breathing than on his heartbeat. This distinction between interoceptive conditions was even more striking when looking at our association results: pain strength was positively correlated with negative emotion, positive emotion and lack of limb ownership when the participant was focusing on his heartbeat, while it was negatively correlated with positive emotion and bodily changes when the participant was focusing on his breathing. These results suggest that perceived pain intensity varies depending on attentional focus, even within interoceptive conditions.

Crucially, the relationship between pain strength and other phenomenal experiences such as emotional states and body perception also seem to vary with the attentional focus. In general, while these conclusions cannot be generalized from a single participant, it appears that for this participant, focusing on his breathing might have a stronger hypoalgesic effect than focusing on the heartbeat.

### The relationship of emotional state and body perception with pain depends on attention

Results from our correlation analyses suggest that emotional states are related to perceived pain intensity, with negative emotion being a good predictor of pain intensity when the participant is focusing on either his painful limb or heartbeat. Pain strength also appears to increase with both perception of bodily changes and lack of limb ownership when the participant is paying attention to his painful limb. However, it might be worth noting that bodily changes were negatively correlated with pain strength in the breathing condition, possibly due to relaxation influencing bodily changes. Overall, these results confirm a strong relationship between chronic pain, emotional states and body distortion in CRPS (Kuttikat et al., 2016). Crucially, this relationship its weaken when attention is diverted from the painful body part. While focusing attention on the painful body part increases perceived pain intensity, on the other hand, focusing on a unaffected body part or breathing seems to have a hypoalgesic effect, in line with previous literature (Bantick et al., 2002; Defina et al., 2021; Eccleston, 1995). Somewhat surprising, focusing on one’s heartbeat seems, at least for this participant, to globally enhance the existing relationship between pain levels and emotional states or body distortion. These results highlight the difference between the interoceptive and exteroceptive conditions (Bantick et al., 2002; Di Lernia, Serino, & Riva, 2016) but also, interestingly, within interoceptive conditions.

### Relationship between power bands and perceived pain intensity

In most of our analyses, power differences did not map onto differences in pain strength when the participant was attending to the painful limb. However, we obtained a positive correlation with pain intensity for the alpha and theta power bands in the unaffected limb condition. These results are interesting as attention to the "other" limb pain ratings can be neurally mapped to these simple neural markers, alpha and theta, but then when attending to other aspects of the body the neural process may be more complex, probably mapping into brain communication, complexity or network measures (e.g., Bullmore & Sporns, 2009; Zhang et al., 2001). While our analysis was not sensitive enough to define the relationship between neurophysiological data and phenomenal experience for the painful limb but show associations for the unaffected limb, future research could look into other neural markers to further investigate the relationship between neural and phenomenal data in chronic pain, such as complexity measures (Lempel-Ziv), heartbeat-evoked potentials (HEP) and EEG frontal asymmetry.

### Future directions

By involving long-term phenomenal and neurophysiological measurements in an ecological setting, this study is the first of its kind. We propose a new method to investigate chronic pain by better sampling phenomenal experience with a pseudo-continuous measure rather than a one-time Likert scale measurement (Jachs et al., 2022). Some of our results highlight the need for new research in the field. The differential impact of interoceptive conditions, as well as the relationship between attentional focus, emotional states and body perception, should be further investigated in future studies on chronic pain. A potential major difference between breathing and heartbeat is that the former can be directly controlled but not the latter, although here both corresponded to passive conditions. Our results might pave the way for a simple neuropsychological intervention which induces pain relief by manipulating interoceptive attention. Future work will aim to generalise those findings across a larger sample of chronic pain patients. We will also aim to identify which subsets of patients might benefit from interoceptive interventions, and which variables predict whether an individual is likely to experience pain relief from interoceptive attention. CRPS and chronic pain is a highly variable condition, and the development of such personalized methods is necessary if we want to better define the relationship between neural and phenomenal data in chronic pain, as well as develop new therapeutic avenues.

## Data Availability

All data produced in the present study are available upon reasonable request to the authors.

## Author note

This research was supported by the Swiss National Science Foundation (grant number: PP00P1_170511 and PP00P1_202674) as well as by the Bertarelli Foundation Catalyst Grant and CCC-LAB internal funds.

Joaquim Streicher and Maria Niedernhuber share first authorship. Bigna Lenggenhager and Tristan Bekinschtein share last authorship.

The authors made the following contributions. Maria Niedernhuber : Conceptualization, Data analysis, Writing - Original Draft Preparation, Writing - Review & Editing; Joaquim Streicher : Conceptualization, Data analysis, Writing - Original Draft Preparation, Writing - Review & Editing; Bigna Lenggenhager : Conceptualization, Writing - Review & Editing, Supervision, Funding acquisition; Tristan Bekinschtein : Conceptualization, Writing - Review & Editing, Supervision, Funding acquisition.

## Acknowledgements

The authors would like to thank Dr Bruno Herbelin, Prof. Olaf Blanke and Arthur Trivier for their support and early contribution. We are indebted to Lavazza and Mokarabia for their continuous support. We are specially thankful for the patient’s commitment and contributions in this first study.

